# Papillary muscles, ventricular loading, and atrial remodelling as beat-to-beat determinants of functional mitral regurgitation: an exploratory Granger causality study

**DOI:** 10.64898/2026.04.03.26350122

**Authors:** Csilla A. Eötvös, Teodora Avram, Eric Daniel Blendea, Matei Ioan Munteanu, Adrian Florin Bubuianu, Madalina P. Moldovan, Petra Hedesiu, Roxana D. Lazar, Iulia G. Zehan, Adriana D. Sarb, Giorgia Coseriu, Patricia Schiop-Tentea, Diana L. Mocan-Hognogi, Roxana Chiorescu, Sorin Pop, Laura Diosan, E. Kevin Heist, Dan Blendea

## Abstract

**Background:** Functional mitral regurgitation results from interacting mechanisms whose contributions vary between atrial and ventricular subtypes and shift within each heartbeat, producing temporal patterns static analyses cannot capture.

**Objectives:** To identify which structural determinants predict mitral regurgitation variability beat to beat using Granger causality within vector autoregression, focusing on papillary muscle dynamics across subtypes.

**Methods:** Frame-level echocardiographic time series from 41 patients (21 atrial, 20 ventricular; 1,959 frames) were z-score standardised within patient. Individual (lag 3) and pooled panel vector autoregression models, with supplementary full-cycle analysis across lags 1–20, tested whether LV volume, left atrial volume, papillary muscle length, and annulus diameter Granger-predict mitral regurgitation area.

**Results:** Individual models revealed marked heterogeneity. In pooled analysis, LV volume was the strongest Granger predictor at the shortest lag (atrial p=0.011; ventricular p=0.006), while left atrial volume reached significance at longer lags (lag 6 atrial p=0.033, ventricular p=0.004). Systolic papillary muscle length was not predictive at any lag. Full-cycle analysis revealed an asymmetric subtype dissociation: papillary muscle length Granger-predicted regurgitation only in the ventricular subtype (lag 1 p<0.001) whereas regurgitation predicted papillary muscle deformation at every lag in the atrial subtype (20/20) but at none in the ventricular subtype. Cross-sectional comparisons of the four determinants did not distinguish MR severity within subtypes.

**Conclusions:** Beat-to-beat vector autoregression and Granger modelling reveals heterogeneous, subtype-specific temporal patterns and a strong directional asymmetry between regurgitation and papillary muscle dynamics across subtypes. This disease-agnostic framework may support patient-specific temporal phenotyping of functional mitral regurgitation.

**Graphical Abstract:** 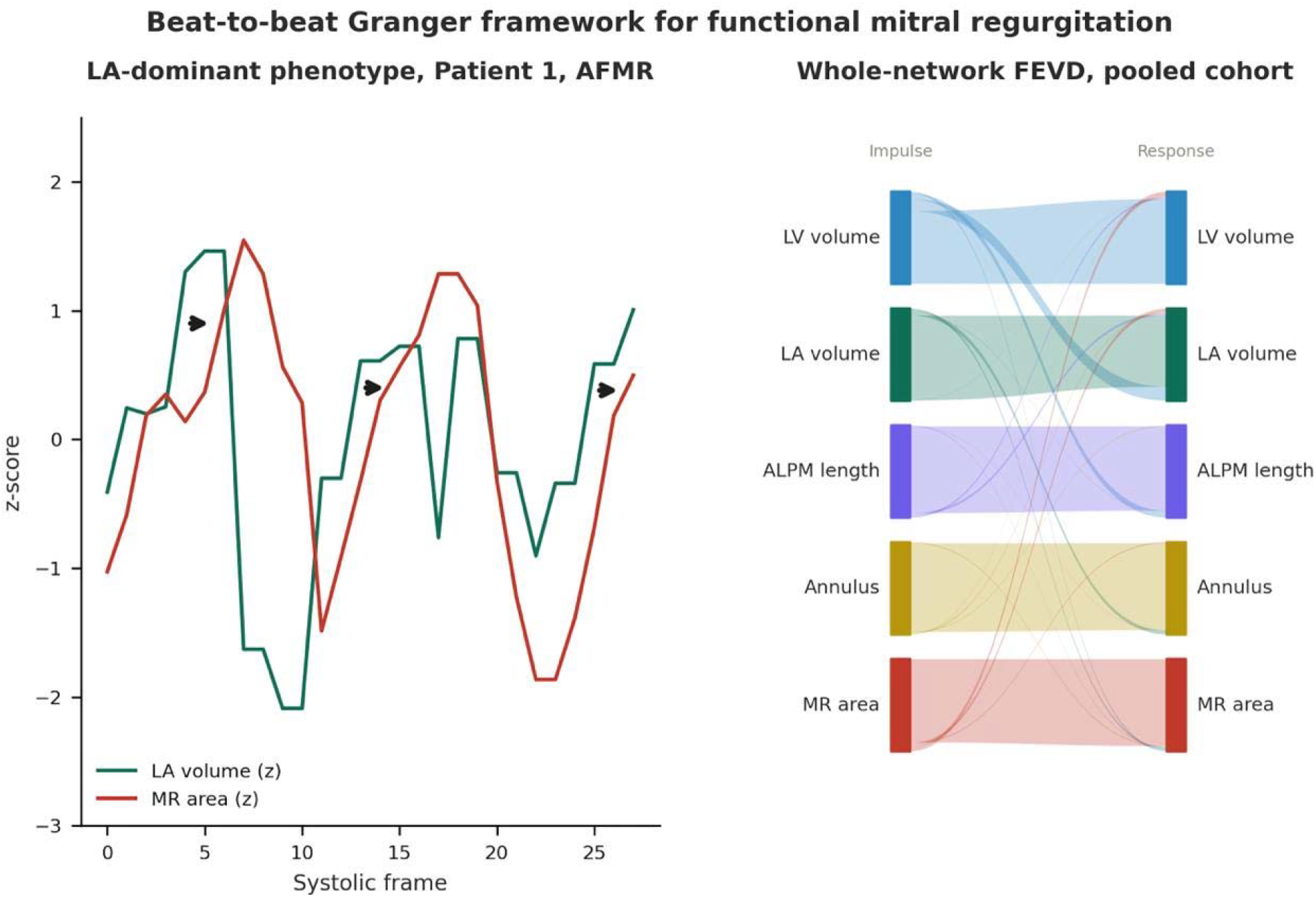

## Introduction

Functional mitral regurgitation (FMR) affects up to 10% of heart failure patients and independently worsens outcomes.^1^ Unlike primary mitral regurgitation (MR), the leaflets are intact; the regurgitation results from ventricular or atrial remodelling that distorts mitral apparatus geometry.^2^ Two subtypes are recognised: ventricular FMR (VFMR), driven by left ventricular (LV) dilatation and apical papillary muscle (PM) displacement; and atrial FMR (AFMR), driven by left atrial (LA) enlargement and annular dilatation in the context of atrial fibrillation or heart failure with preserved ejection fraction.^3, 4^

A subset responds inadequately to treatment, possibly reflecting mechanistic heterogeneity that current interventional strategies fail to address and may sometimes exacerbate (Figure 1-Panel A).^5^ Functional MR represents the common endpoint of multiple interacting pathways: ventricular volume overload, annular dilatation, leaflet tethering, and PM displacement, whose relative contributions vary substantially between patients.^6, 7^ COAPT^8^ and MITRA-FR^9^ illustrate this heterogeneity: morphologically similar patients may have fundamentally different dominant drivers and respond differently to the same intervention. A therapy targeting annular geometry will fail if the primary driver is PM tethering. Identifying the dominant mechanism individually remains an unresolved challenge.

Papillary muscles (PMs) are mechanistically central to FMR but clinically undercharacterised. In VFMR, PM displacement increases leaflet tethering in post-infarction MR and dilated cardiomyopathy.^10-13^

Two questions remain. First, whether PM displacement actively modulates beat-to-beat MR variability or merely establishes a fixed tethering substrate; only in the former case would PM-targeted therapy reduce dynamic MR fluctuation. Second, whether PMs contribute dynamically to MR in AFMR.

Static cross-sectional measurements capture geometric correlates of MR severity but cannot establish directionality: the same correlation may reflect tethering causing MR, remodelling causing both, or MR-driven overload displacing the PMs. Disentangling these relationships requires a framework that accounts for temporal precedence and quantifies each determinant’s independent contribution. Granger causality within vector autoregressive (VAR) modelling of beat-to-beat time series provides exactly this, and is facilitated by the substantial beat-to-beat variability of MR ^14-16^ that supplies the signal variation such models require. Developed in econometrics ^17^, and applied in cardiovascular physiology^18,19^ Granger causality tests whether a variable’s past improves prediction of another beyond its own history. Throughout, ‘causality’ denotes temporal predictability, not mechanistic causation (Figure 1 – Panel B).

We hypothesized that beat-to-beat MR variability reflects temporally structured interactions among ventricular loading, atrial remodelling, annular geometry, and PM dynamics, differing between MR subtypes.

## Methods

### Study population and echocardiographic data acquisition

We retrospectively analysed transthoracic echocardiograms from 41 patients (21 AFMR, 20 VFMR) enrolled at three university hospitals in Cluj County, Romania (Figure 2); 22 patients (19 AFMR, 3 VFMR) were in atrial fibrillation at the time of examination. Ventricular FMR required LV dilatation or dysfunction with intact leaflets; AFMR required LA enlargement with preserved or mildly reduced LV function without significant ventricular remodelling. Measurements were performed according to the ASE/EACVI guidelines^20^. Baseline structural variables included PM length, tenting height, and mitral annulus diameter (Figure 3). The anterolateral PM (ALPM) length was measured in the apical four-chamber view from the ALPM apex to the mid-basal papillary muscle insertion (Figure 3).^13^ Per patient, two or more consecutive cycles were analysed frame-by-frame at 30 fps (Figure 1 – Panel C), yielding 21–181 systolic frames per patient (median 29; 1,959 frames total across all 41 patients, systolic and diastolic combined). Lag-to-time mapping is approximate: at 30 fps a single lag corresponds to ≈33 ms, but in the systolic-frames-only pooled VAR consecutive frames are not separated by a uniform interval, and even short lags may straddle beat boundaries depending on heart rate. Lag values throughout the manuscript are therefore reported as analytic units rather than as fixed within-beat or cross-beat intervals. Measurements were performed by three observers (one per patient).

### Variable selection

Four determinants were selected for MR variability analysis: LV volume and LA volume (ventricular loading and atrial remodelling), together with mitral annulus diameter and ALPM length measured in systolic frames (annular and subvalvular geometry).

### Frame-by-frame longitudinal measurements

Five variables were measured every frame: MR jet area (cm^2^), LV volume (ml), LA volume (ml), annulus diameter (cm), and ALPM length (mm). Tenting height was measured in systolic frames only. Frame-by-frame LV and LA volumes were calculated using the biplane area-length method, recommended by the ASE/EACVI guidelines^20^as an acceptable alternative when disk summation is impractical. Mitral annulus diameter was tracked continuously across the cardiac cycle in the apical four-chamber view as the septal-to-lateral annular distance, a plane validated for mitral annulus measurement in FMR. ^21^Mitral valve tenting height was tracked in systolic frames only in the apical four-chamber view as the perpendicular distance from the mitral annular plane to the leaflet coaptation point; the apical four-chamber view has been identified as the preferred plane for tenting height quantification because the annular plane is most clearly defined in this view^2^, and frame-by-frame tracking of tenting dynamics in functional MR has been previously reported.^22^The VAR/Granger framework isolates within-patient temporal changes from each patient’s own mean, so any systematic measurement offset arising from the area-length method, the apical four-chamber annular plane, or tenting-height landmark placement shifts that mean but does not affect the estimated lag structure or directional dependencies between variables. Frames were classified as systolic or diastolic. Mitral regurgitant jet area served as a temporal variability surrogate, not an absolute severity metric. Quantitative approaches (EROA, PISA, continuity equation) and vena contracta width are preferred for grading MR severity^23^but are not feasible frame-by-frame: the quantitative methods rely on multi-step calculations and assumptions that are not stable across frames, and vena contracta width (a few millimetres) falls below the spatial resolution needed for reliable tracking at 30 fps. Jet area, despite its load dependence, is the only descriptor available at every frame, and within-patient relative fluctuations remain informative. All diastolic frames were retained in the time series. MR area was recorded as zero in frames without regurgitant flow and as the measured jet area in frames with diastolic regurgitation.

### Within-patient z-score standardisation

All time-series variables were z-score standardised within patient (subtracting each patient’s mean and dividing by their standard deviation) to remove between-subject magnitude differences while preserving temporal variability. Variables spanning the full cardiac cycle used all frames; MR area was z-scored within systolic frames only, since including zero-valued diastolic frames would distort the within-patient standardisation.

### Statistical analysis

All analyses are exploratory and intended to demonstrate an analytical framework rather than to establish definitive findings about FMR subtypes. Baseline characteristics are summarised as median (interquartile range) for continuous variables and count (percentage) for categorical variables, given the small sample size and visibly skewed distributions of several variables. Between-group comparisons used the Mann-Whitney U test for continuous variables and Fisher exact test for categorical variables. No correction for multiple comparisons was applied.

### Individual-patient vector autoregression

A VAR model was fitted per patient (minimum 20 systolic frames), expressing each variable as a function of its own and all other variables’ lagged values. Endogenous variables were MR area, LV volume, LA volume, and ALPM length (three-variable model when ALPM was not visible). With short series (median 29 frames), individual models primarily illustrate heterogeneity. Lag orders of 1 to 3 frames (∼33–100 ms at 30 fps) were fitted per patient, with lag 3 as the primary analysis and lags 1 and 2 as sensitivity analyses. Stability was verified by eigenvalue analysis.

### Pooled panel vector autoregression

A pooled panel VAR was estimated separately for AFMR, VFMR, and the combined cohort, using systolic frames from patients with ALPM visibility. Five endogenous variables were included: MR area, LV volume, LA volume, systolic ALPM length, and annulus diameter. Each patient had a separate baseline absorbed into the model. Models were fitted at lag orders 2, 6, and 7, with lag 2 as the primary specification and lags 6 and 7 as robustness checks. The global systolic frame counter served as the time variable. Granger causality, FEVD, and OIRF were extracted as for individual models.^24^All analyses were performed in Stata 19 (StataCorp, College Station, TX). A two-sided p<0.05 was considered significant throughout.

### Full-cycle pooled vector autoregression

To test whether PM dynamics across the full cardiac cycle contribute to MR prediction, a supplementary pooled panel VAR was estimated separately for AFMR and VFMR. This model differed from the primary systolic pooled VAR in three respects: it included four endogenous variables rather than five (MR area, LV volume, LA volume, and ALPM length; annulus diameter excluded because it is measured only in systolic frames); variables were z-score standardised across the full cardiac cycle rather than within systolic frames only; and lag orders 1 through 20 were each fitted, in both forward and reverse directions, with FEVD evaluated at horizon 10. Full per-lag results appear in Supplementary Table S1.

## Results

### Baseline Characteristics

Atrial FMR patients were older, predominantly female, and more often in atrial fibrillation; VFMR patients were predominantly male with more coronary disease (Table 1). In VFMR, moderate–severe MR was significantly associated with more frequent LBBB and greater early-systolic tenting height, with non-significant trends in the same direction for lower ejection fraction, higher tricuspid regurgitation grade, and higher NYHA class. In AFMR, only tricuspid regurgitation grade was significantly associated with MR severity, and LV volume and ejection fraction were preserved. LV end diastolic volume was modestly higher in moderate–severe MR in the overall cohort, driven by the cross-subtype distribution rather than within-subtype severity gradients. None of the four VAR determinants (LV volume, LA volume, end-systolic annulus diameter, ALPM length) differed significantly by MR severity within either subtype; these cross-sectional comparisons are limited by subgroup size and are reported for completeness, as the primary analysis focuses on beat-to-beat temporal dynamics.

**Table 1.** Baseline characteristics by MR severity — mild vs moderate–severe.

| Variable | AFMR (n=21) |  |  | VFMR (n=20) |  |  | Overall (n=41) |  |  |
| --- | --- | --- | --- | --- | --- | --- | --- | --- | --- |
|  | Mild (n=10) | Mod-Sev (n=11) | p | Mild (n=8) | Mod-Sev (n=12) | p | Mild (n=18) | Mod-Sev (n=23) | p |
| **Demographics** |  |  |  |  |  |  |  |  |  |
| **Age (years)** | 83.0 (76.0–86.0) | 78.0 (77.0–83.0) | 0.378 | 64.0 (56.0–68.5) | 68.0 (67.5–73.0) | 0.112 | 74.0 (65.0–84.0) | 75.0 (68.0–79.0) | 0.875 |
| **Female sex** | 8 (80%) | 8 (73%) | 1.000 | 1 (12%) | 2 (17%) | 1.000 | 9 (50%) | 10 (43%) | 0.758 |
| **CAD** | 2 (20%) | 0 (0%) | 0.214 | 6 (75%) | 8 (67%) | 1.000 | 8 (44%) | 8 (35%) | 0.748 |
| **AF history** | 8 (80%) | 9 (82%) | 1.000 | 2 (25%) | 5 (42%) | 0.642 | 10 (56%) | 14 (61%) | 0.760 |
| **NYHA class** | 2.5 (2.0–3.0) | 2.0 (2.0–3.0) | 0.226 | 2.0 (2.0–2.5) | 3.0 (2.0–3.0) | 0.068 | 2.0 (2.0–3.0) | 2.0 (2.0–3.0) | 0.622 |
| **ECG** |  |  |  |  |  |  |  |  |  |
| **LBBB** | 2 (20%) | 3 (27%) | 1.000 | 0 (0%) | 6 (50%) | **0.042** | 2 (11%) | 9 (39%) | 0.075 |
| **Echocardiography** |  |  |  |  |  |  |  |  |  |
| **MR grade (1–4)** | 1.0 (1.0–1.0) | 3.0 (2.0–4.0) | **<0.001** | 1.0 (1.0–1.0) | 2.5 (2.0–3.0) | **<0.001** | 1.0 (1.0–1.0) | 3.0 (2.0–3.0) | **<0.001** |
| **TR grade** | 1.0 (1.0–2.0) | 2.0 (2.0–2.0) | 0.036 | 1.0 (0.5–1.0) | 1.0 (1.0–2.0) | 0.054 | 1.0 (1.0–2.0) | 2.0 (1.0–2.0) | **0.012** |
| **Pulmonary hypertension** | 5 (50%) | 8 (73%) | 0.387 | 1 (12%) | 3 (25%) | 0.762 | 6 (33%) | 11 (48%) | 0.426 |
| **LVEDV (ml)** | 85.5 (70.0–98.0) | 102 (81–107) | 0.067 | 121 (110–158) | 148 (128–294) | 0.090 | 98.5 (71.0–126.0) | 127 (100–205) | **0.047** |
| **Ejection fraction (%)** | 56.6 (52.7–61.0) | 54.7 (50.5–56.8) | 0.342 | 42.0 (40.0–48.0) | 31.0 (22.5–42.5) | 0.057 | 52.6 (44.0–58.2) | 50.0 (30.0–54.7) | 0.144 |
| **LA volume (ml)** | 93.0 (76.0–114.0) | 101 (73–185) | 0.418 | 78.5 (49.5–98.0) | 63.0 (46.5–77.0) | 0.375 | 87.0 (72.0–102.0) | 75.0 (56.0–104.0) | 0.684 |
|  | Mild (n=10) | Mod-Sev (n=11) | p | Mild (n=8) | Mod-Sev (n=12) | p | Mild (n=18) | Mod-Sev (n=23) | p |
| MV annulus end-systole (cm) | 2.5 (2.2–3.0) | 2.9 (2.5–3.2) | 0.290 | 3.1 (2.6–3.3) | 3.2 (2.5–3.4) | 0.908 | 2.8 (2.3–3.2) | 2.9 (2.5–3.3) | 0.423 |
| Tenting height early systole (cm) | 0.9 (0.8–1.2) | 1.1 (0.6–1.3) | 0.622 | 0.9 (0.9–1.0) | 1.2 (1.0–1.4) | **0.034** | 0.9 (0.9–1.1) | 1.2 (0.9–1.3) | 0.070 |
| ALPM length (cm) | 2.6 (2.5–3.1) | 2.4 (2.1–2.6) | 0.067 | 2.7 (2.2–3.2) | 2.9 (2.4–3.5) | 0.700 | 2.7 (2.4–3.1) | 2.5 (2.2–3.0) | 0.438 |

1 Data presented as median (interquartile range) for continuous variables and n (%) for categorical variables. p values from Mann–Whitney U 2 test (continuous) or Fisher exact test (categorical). Bold p values indicate p<0.05. Mild = MR grade 1; Moderate–Severe = MR grade ≥2. AFMR 3 = atrial functional mitral regurgitation; VFMR = ventricular functional mitral regurgitation; CAD = coronary artery disease; AF = atrial fibrillation; NYHA = New York Heart Association; LBBB = left bundle branch block; TR = tricuspid regurgitation; LVEDV = LV end-diastolic volume; LA = left atrium; MV = mitral valve; ALPM = anterolateral papillary muscle.

### Individual Patient VAR Analysis

Patients with sufficient systolic frames underwent individual VAR analysis (Figure 4, Table 2). The primary analysis used fixed lag 3 (∼100 ms), the longest lag compatible with the shortest series, with lags 1 and 2 as directionally consistent sensitivity analyses. Of 34 analysable patients, 25 had stable models at lag 3; patients without ALPM visibility used a reduced three-variable model.

**Table 2.**
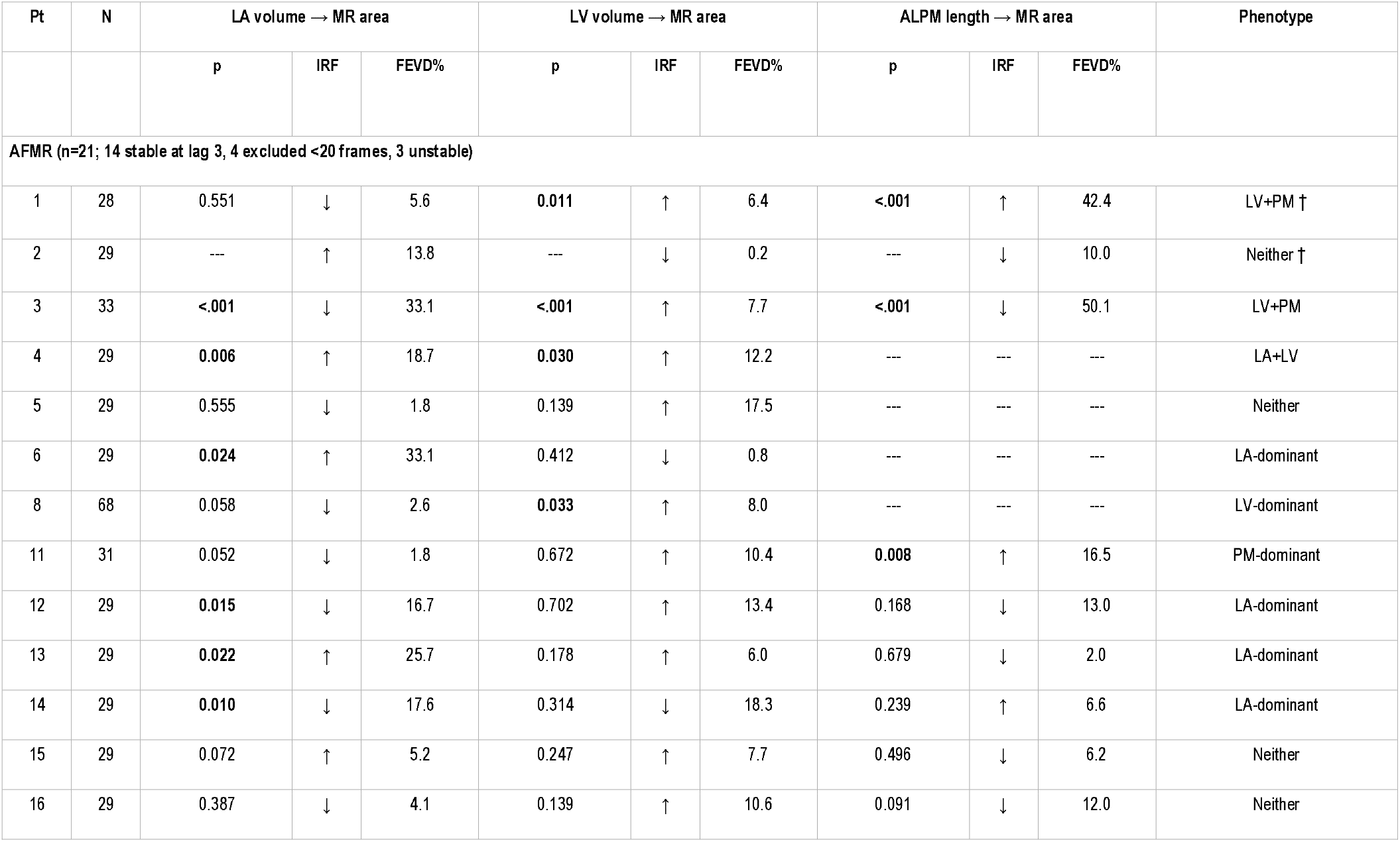
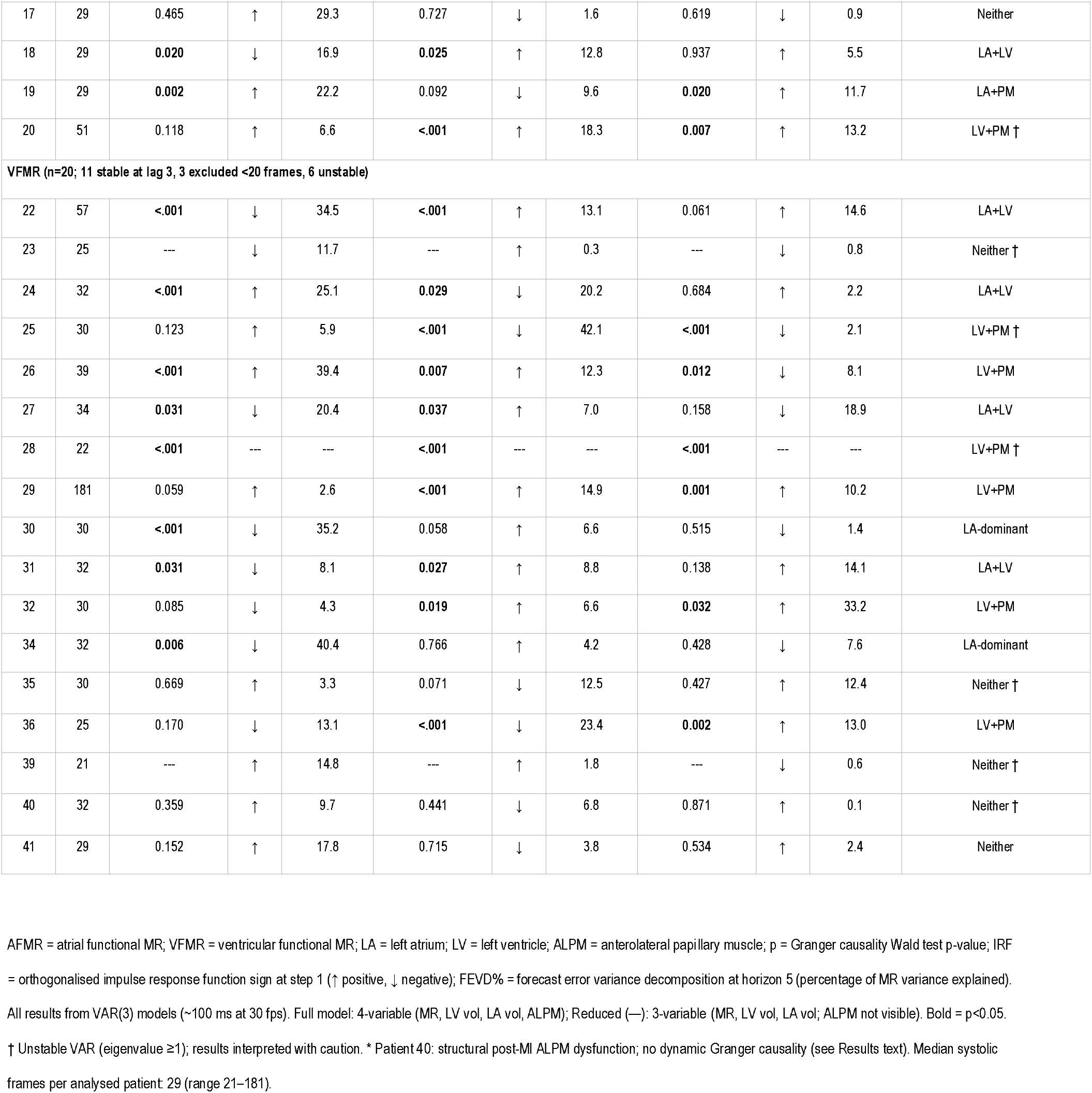
Individual Patient VAR Granger Causality Results.

The most common phenotype in both subtypes was “neither,” reflecting limited power with short series; individual phenotypes should therefore be interpreted as illustrative rather than definitive. LV and LA involvement were equally represented in both subtypes, whereas papillary muscle involvement was more frequent in VFMR (6 of 10 patients with PM-containing phenotypes; 4 of 6 among stable models), suggesting that active beat-to-beat PM–MR coupling is primarily ventricular in this cohort. Patient 40 (VFMR) illustrates this distinction: despite post-infarction PM dysfunction and elevated tenting, no Granger predictors were identified, showing that fixed geometric deformation does not necessarily translate into active dynamic PM–MR coupling.

### Measurement Reproducibility

Intraobserver reproducibility (7 patients, 70 paired observations) yielded excellent ICC for MR area (0.986), LA volume (0.981), and LV volume (0.971), and a good ICC for ALPM length (0.890). Mean bias was negligible across all variables; detailed Bland–Altman results are provided in Supplementary Table S2.

### Pooled VAR Analysis

The pooled VAR included systolic frames from 37 patients with ALPM visibility. After lag initialisation, 463 AFMR, 704 VFMR, and 1,167 overall observations remained; all models were stable (Table 3).

**Table 3.** Pooled VAR Granger Causality — Beat-to-Beat Predictors of MR Area.

|  |  | AFMR (17 pts) |  |  | VFMR (20 pts) |  |  | Overall (37 pts) |  |  |
| --- | --- | --- | --- | --- | --- | --- | --- | --- | --- | --- |
| Phase | Lag | p | IRF | FEVD | p | IRF | FEVD | p | IRF | FEVD |
| LA volume → MR area |  |  |  |  |  |  |  |  |  |  |
| Systolic | 2 | 0.509 |  | 0.0 | 0.018 | + | 0.7 | <0.001 | + | 0.8 |
| Systolic | 6 | 0.033 | - | 0.5 | 0.004 | + | 1.1 | <0.001 | + | 1.1 |
| Systolic | 7 | 0.043 | - | 0.5 | 0.011 | + | 1.2 | <0.001 | + | 1.1 |
| LV volume → MR area |  |  |  |  |  |  |  |  |  |  |
| Systolic | 2 | 0.011 | + | 2.1 | 0.006 | + | 1.9 | <0.001 | + | 2.2 |
| Systolic | 6 | 0.012 | + | 2.3 | 0.328 |  | 1.0 | 0.009 | + | 1.8 |
| Systolic | 7 | 0.047 | + | 1.5 | 0.313 |  | 0.9 | 0.044 | + | 1.3 |
| ALPM length → MR area |  |  |  |  |  |  |  |  |  |  |
| Systolic | 2 | 0.457 | - | 0.5 | 0.831 |  | 0.2 | 0.977 |  | 0.01 |
| Systolic | 6 | 0.295 |  | 1.0 | 0.819 |  | 0.02 | 0.884 |  | 0.1 |
| Systolic | 7 | 0.210 |  | 1.2 | 0.707 |  | 0.03 | 0.243 |  | 0.1 |
| Annulus diameter → MR area |  |  |  |  |  |  |  |  |  |  |
| Systolic | 2 | 0.966 | - | 0.0 | 0.238 | - | 0.6 | 0.116 | - | 0.5 |
| Systolic | 6 | 0.033 | - | 0.2 | 0.017 | - | 0.03 | 0.436 |  | 0.3 |
| Systolic | 7 | 0.027 | - | 0.3 | 0.011 | + | 0.06 | 0.349 |  | 0.2 |
| ALL predictors (joint Granger test) |  |  |  |  |  |  |  |  |  |  |
| Systolic | 2 | 0.252 | — | — | 0.029 | — | — | <0.001 | — | — |
| ALPM length → MR area (full-cycle model ‡) |  |  |  |  |  |  |  |  |  |  |
| Full cycle | 1 | 0.314 |  | 0.3 | <0.001 | + | 1.3 | — |  |  |
| Full cycle | 2 | 0.544 |  | 0.4 | 0.005 | + | 1.3 | — |  |  |
| Full cycle | 4 | 0.502 |  | 0.5 | 0.030 | + | 1.4 | — |  |  |
| Full cycle | 7 | 0.411 |  | 0.9 | 0.040 | + | 1.5 | — |  |  |
| MR area → ALPM length (reverse Granger, full-cycle model ‡) |  |  |  |  |  |  |  |  |  |  |
| Full cycle | 1 | 0.016 |  | 1.4 | 0.289 |  | 2.2 | — |  |  |
| Full cycle | 2 | <0.001 |  | 3.8 | 0.359 |  | 2.0 | — |  |  |
| Full cycle | 4 | <0.001 |  | 6.4 | 0.183 |  | 3.0 | — |  |  |
| Full cycle | 7 | <0.001 |  | 6.7 | 0.621 |  | 2.5 | — |  |  |

Pooled VAR with patient fixed effects; 5 endogenous variables (MR area, LV volume, LA volume, systolic ALPM length, annulus diameter); all models stable. Lag 2 ≈ 67 ms, lag 6 ≈ 200 ms, and lag 7 ≈ 233 ms, at 30 fps acquisition; lag-to-time mapping is approximate (see Methods). p = Granger causality Wald $\chi^2$ test; bold = p < 0.05. IRF = impulse response sign at step 1 (shown only for significant predictors). FEVD = forecast error variance decomposition at horizon 5 (% of MR area variance). † 4 AFMR patients excluded (absent ALPM). ‡ Full-cycle rows: 4-variable model (MR area, LV volume, LA volume, ALPM length); z-scores standardised across full cardiac cycle; FEVD at horizon 10.

Left ventricular volume was the strongest individual Granger predictor of MR area in both subtypes (AFMR p=0.011; VFMR p=0.006), whereas systolic ALPM length was not predictive at any of the three lags examined — lag 2 (∼67 ms), lag 6 (∼200 ms), or lag 7 (∼233 ms) — with FEVD contributions consistently below 1.5%. The joint test of all four predictors was significant in VFMR (p=0.029) and overall (p<0.001) but not in AFMR (p=0.252), likely reflecting the limited power of the smaller AFMR sample (463 observations, 17 patients) spread across eight degrees of freedom rather than absence of multifactorial coupling. These findings together suggest that PM deformation primarily establishes the structural tethering substrate rather than actively driving beat-to-beat MR variability, consistent with the rarity of purely PM-dominant phenotypes in the individual analysis.

### Full-cycle papillary muscle analysis

To test whether PM dynamics across the cardiac cycle contribute to MR prediction, a supplementary full-cycle VAR analysis was performed. Multivariate models including MR area, LA volume, LV volume, and ALPM length (with patient fixed effects) were fitted at lags 1–20 in each subtype (17 AFMR, 20 VFMR patients; full per-lag results in Supplementary Table S1). Anterolateral PM Granger-predicted MR only in VFMR, primarily at lags 1 through 4 (lag 1 p<0.001) and at lag 7 (p=0.040), with FEVD contributions of 1.3–1.5% (Table 3). In AFMR, ALPM was non-predictive at every lag (0 of 20 significant). The reverse direction showed an asymmetric pattern: MR predicted ALPM at all 20 lags in AFMR (lag 1 p=0.016, lags 2–20 all p<0.001) but at no lag in VFMR (minimum p=0.061 at lag 9). Consistent with this, ALPM variance was 66–72% self-driven in AFMR (LV volume contributing 17–20%, LA volume 6–9%), compared with 95–96% self-driven in VFMR.

### Lag-specific temporal windows

The three lag windows revealed a progressive shift in the dominant predictor. At lag 2, LV volume was the strongest predictor in both subtypes (AFMR p=0.011; VFMR p=0.006), with no detectable contribution from ALPM. By lag 6, the picture diverged: LA volume became significant in both subtypes (AFMR p=0.033; VFMR p=0.004), annulus diameter became significant in VFMR (lag 6 p=0.017, lag 7 p=0.011), and LV volume lost predictive value in VFMR (p=0.328) while persisting in AFMR (p=0.012). LA volume remained significant in AFMR at lag 7 as well (p=0.043). Systolic ALPM remained non-predictive across all three lags in both subtypes (Table 3). Together, these patterns suggest that ventricular loading dominates the shortest lag whereas atrial remodelling and annular dynamics emerge as predictors at longer lags.

Whole-network forecast error variance decomposition (Figure 5) confirmed this architecture: MR area was overwhelmingly self-driven (AFMR 97.3%, VFMR 96.6%), with LV volume and LA volume as the largest external contributors, and ALPM length explaining less than 1% of MR variance in either subtype.

## Discussion

To our knowledge, this is the first application of beat-to-beat vector autoregression and Granger causality to frame-level echocardiography in FMR. Functional MR determinants interact dynamically across multiple timescales; given the exploratory design, these findings describe temporal predictive patterns rather than stable mechanistic phenotypes. The framework is disease-agnostic and transferable to other conditions with dynamic echocardiographic data.

### Individual phenotypic heterogeneity

Most individual patients showed no dominant Granger predictor, yet pooled analysis consistently identified LV volume as a driver of MR variability in both subtypes, distributed multifactorial coupling at the individual level resolving into a coherent signal only when variance is pooled. Among identifiable phenotypes, LV and LA involvement were equally represented in both subtypes, whereas PM involvement appeared predominantly in VFMR.

The PM role differed sharply between subtypes. In VFMR, ALPM predicted MR at short lags, with the diastole-to-systole transition carrying predictive information that a systolic-only analysis truncates. The variance contribution is modest but independent of ventricular loading. In AFMR, the relationship ran in the opposite direction: regurgitation predicted PM deformation at every lag, and the forward direction never reached significance. The PM appears to establish a tethering substrate on which ventricular loading operates. Papillary muscle targeted therapy could modify this substrate in VFMR.

### Systolic ALPM and lag-specific temporal patterns

Different lag orders revealed how each determinant acts on MR over time. Systolic ALPM length was not predictive at any of the three lags examined, consistent with a fixed tethering substrate rather than beat-by-beat contribution. LV volume was strongest at the shortest lag but lost predictive value at longer lags in VFMR, while LA volume reached significance in AFMR only at longer lags, indicating that ventricular loading and atrial remodelling act on different temporal scales.

Asking whether MR predicts ALPM clarified the direction of influence. Mitral regurgitation predicted ALPM deformation at every lag in AFMR but at no lag in VFMR. In AFMR the relationship runs almost entirely from regurgitation to ALPM length, consistent with PM length change being a downstream consequence of volume overload. In VFMR, no reverse pattern emerges, which together with the short-lag forward coupling indicates that PM geometry contributes modestly but independently to MR variability.

Variance decomposition (Figure 5) showed that MR was overwhelmingly self-driven in both subtypes, with LV and LA volume as the largest external contributors. ALPM accounted for a minimal proportion, reinforcing the tethering-substrate interpretation.

### Clinical implications and the COAPT/MITRA-FR paradox

The subtype-specific temporal patterns reported here may partly explain the divergent outcomes of COAPT^8^and MITRA-FR^9^: patients with similar static geometry may differ in how regurgitation couples with ventricular loading, annular dilatation, and PM position from beat to beat. Where ventricular loading dominates variability, remodelling-targeted therapies may be preferred; where tethering predominates, interventions directed at the PM or the mitral apparatus may be more effective, given that PM deformation contributes modestly but independently to MR in VFMR. ^25, 26^ Temporal phenotyping could therefore complement static morphology in trial stratification, pending prospective validation in independent cohorts.

Beat-to-beat VAR/Granger modelling reveals subtype-specific temporal signatures of FMR and a directional asymmetry between regurgitation and PM dynamics. These findings are hypothesis-generating and require prospective validation.

## Limitations

The analysis is exploratory: Granger “causality” denotes temporal predictive content, not mechanistic causation, and no multiple-comparisons correction was applied. Individual time series were short (21–181 systolic frames; median 29), limiting power and producing frequent “neither” phenotypes. Patients were enrolled at three hospitals but selected for adequate image quality, so prospective multicentre replication is essential; measurements were performed by three observers (one per patient), with intraobserver reproducibility in Supplementary Table S2. MR jet area was used as a high-temporal-resolution surrogate; within-patient fluctuations remain informative despite its load dependence. Individual models used fixed lag 3 with lags 1 and 2 as sensitivity analyses, and the pooled panel VAR assumes a homogeneous lag structure, with robustness confirmed across lags 2, 6, and 7. Longitudinal analysis was restricted to the ALPM. In the pooled model, lagged observations could in principle cross patient boundaries, but such transitions are negligible and are unlikely to influence results. Finally, the VAR/Granger framework captures only linear temporal dependencies; nonlinear interactions are explored in a companion study.

## Abbreviations

AFMR: atrial functional mitral regurgitation
ALPM: anterolateral papillary muscle
FEVD: Forecast error variance decomposition
FMR: functional mitral regurgitation
LA: left atrium
LV: left ventricle
MR: mitral regurgitation
OIRF: Orthogonalised impulse response function
PM: papillary muscle
VAR: vector autoregressive
VFMR: ventricular functional mitral regurgitation.

## Legends

**Figure 1.** Panel A: Conceptual framework of beat-to-beat Granger-predictive modelling in FMR. The determinant network shows how MR jet area variability is influenced by LV volume, LA volume, annulus diameter, and ALPM length within each cardiac cycle. Bidirectional arrows indicate tested Granger-predictive pathways; Panel B: Temporal logic of Granger causality testing. Past values of variable X at lags t−2 and t−1 are tested as predictors of variable Y at time t, conditional on Y’s own history. If including past values of X improves prediction of Y beyond Y’s autoregressive component, X is said to Granger-cause Y. In this study, X represents each structural determinant (LV volume, LA volume, ALPM length, annulus diameter) and Y represents MR area. Panel C: Schematic of frame-by-frame echocardiographic acquisition showing systolic frame labelling across consecutive cardiac cycles. ALPM = anterolateral papillary muscle; FMR = functional MR; LA = left atrium; LV = left ventricle; MR = mitral regurgitation.

**Figure 2.** Study flow, phenotype classification, and VAR analysis framework.

**Figure 3.** Four-chamber apical view illustrating key structural measurements of the mitral valve apparatus. The mitral annulus (green line) and tenting height (yellow line) are shown, along with the endocardial contour (dotted line). Anterolateral papillary muscle (ALPM) length (blue line) was measured as the distance from the LV apex to its mid-basal insertion along the endocardial border.

**Figure 4.** Individual-patient Granger-predictive phenotypes. Each patient is classified by the dominant frame-level Granger predictor of MR jet area (LV volume, LA volume, or papillary muscle length). Classification based on Wald FEVD at horizon 5. Phenotype labels correspond to Table 2. AFMR = atrial functional MR; VFMR = ventricular functional MR

**Figure 5.** Whole-network forecast error variance decomposition in AFMR (A, B) and VFMR (C, D). Pooled VAR(2) with patient fixed effects, FEVD at horizon 5, upstream-first Cholesky ordering. Ribbon width represents the percentage of forecast variance in the response variable explained by the impulse variable. Bar charts show the MR area variance breakdown for each subtype.

## Ethics statement. Consent

This retrospective study was conducted in accordance with the Declaration of Helsinki. Institutional review board approval was obtained from the Ethics Committee of Iuliu Haţieganu University of Medicine and Pharmacy, Cluj-Napoca, Romania (approval number 216, 20 July 2022). Written informed consent was obtained from all participants prior to enrolment. Data from 41 consecutive patients with functional mitral regurgitation were retrospectively analysed.

## Data availability

The de-identified patient-level dataset is archived on Zenodo (10.5281/zenodo.19396466) and available upon reasonable request to the corresponding author, subject to a data use agreement. The Stata analysis code is archived on Zenodo (10.5281/zenodo.19494114) and freely available at https://github.com/dblendea/FMR_causality.

A preprint version of this manuscript is available at https://medrxiv.org/cgi/content/short/2026.04.03.26350122v1.

## Author contributions

All authors contributed to the conception and design of the study, interpretation of data, drafting and critical revision of the manuscript, and approved the final version for submission. D Blendea and CA Eotvos had primary responsibility for the final content.

