## Supplemental Tables for "Papillary muscles, ventricular loading, and atrial remodelling as beat-to-beat determinants of functional mitral regurgitation: an exploratory Granger causality study"

**Supplementary Table S1.** Full-cycle pooled VAR Granger causality — per-lag p-values for the 20-lag sweep

| Lag | AFMR (17 patients, 617 observations) | | | | VFMR (20 patients, 1,186 observations) | | | |
|---|---|---|---|---|---|---|---|---|
| | ALPM→MR | MR→ALPM | LV→MR | LA→MR | ALPM→MR | MR→ALPM | LV→MR | LA→MR |
| 1 | 0.314 | **0.016** | **0.003** | 0.440 | **0.001** | 0.288 | **<0.001** | 0.319 |
| 2 | 0.544 | **<0.001** | **0.029** | 0.642 | **0.005** | 0.359 | **<0.001** | 0.052 |
| 3 | 0.539 | **<0.001** | **0.001** | 0.474 | **0.010** | 0.441 | **<0.001** | **0.009** |
| 4 | 0.502 | **<0.001** | **0.001** | 0.604 | **0.030** | 0.183 | **<0.001** | **0.024** |
| 5 | 0.690 | **<0.001** | **0.002** | 0.365 | 0.056 | 0.363 | **<0.001** | **0.011** |
| 6 | 0.657 | **<0.001** | **0.005** | 0.486 | 0.052 | 0.487 | **0.002** | **0.012** |
| 7 | 0.411 | **<0.001** | **0.024** | 0.145 | **0.040** | 0.621 | **0.011** | **0.019** |
| 8 | 0.400 | **<0.001** | **0.035** | 0.074 | 0.059 | 0.673 | **0.022** | **0.029** |
| 9 | 0.412 | **<0.001** | 0.055 | **0.033** | 0.063 | 0.061 | 0.065 | **0.031** |
| 10 | 0.369 | **<0.001** | 0.111 | **0.043** | 0.089 | 0.065 | 0.098 | 0.051 |
| 11 | 0.237 | **<0.001** | 0.076 | **0.036** | 0.092 | 0.097 | 0.168 | **0.018** |
| 12 | 0.268 | **<0.001** | 0.178 | **0.006** | 0.132 | 0.100 | 0.105 | **0.028** |
| 13 | 0.251 | **<0.001** | 0.082 | **0.009** | **0.035** | 0.134 | 0.160 | **0.025** |
| 14 | 0.314 | **<0.001** | 0.053 | **0.017** | 0.050 | 0.140 | 0.126 | **0.014** |

| Lag | AFMR (17 patients, 617 observations) | | | | VFMR (20 patients, 1,186 observations) | | | |
|---|---|---|---|---|---|---|---|---|
| | **ALPM→MR** | **MR→ALPM** | **LV→MR** | **LA→MR** | **ALPM→MR** | **MR→ALPM** | **LV→MR** | **LA→MR** |
| 15 | 0.265 | **<0.001** | 0.124 | **0.010** | 0.069 | 0.117 | 0.090 | 0.053 |
| 16 | 0.258 | **<0.001** | **0.013** | **<0.001** | 0.059 | 0.123 | 0.114 | **0.004** |
| 17 | 0.349 | **<0.001** | **0.003** | **0.001** | 0.053 | 0.150 | 0.128 | **<0.001** |
| 18 | 0.121 | **<0.001** | **<0.001** | **<0.001** | 0.074 | 0.180 | 0.171 | **0.003** |
| 19 | 0.104 | **<0.001** | **<0.001** | **<0.001** | **0.040** | 0.153 | 0.218 | **0.006** |
| 20 | 0.107 | **<0.001** | **<0.001** | **<0.001** | 0.053 | 0.065 | 0.238 | **0.009** |

*Pooled panel VAR with patient fixed effects, fitted separately for AFMR and VFMR, using z-scored full-cycle observations (MR area, LA volume, LV volume, ALPM length). For each lag order (1 to 20), four Granger causality tests are reported: ALPM length → MR area (forward), MR area → ALPM length (reverse), LV volume → MR area, and LA volume → MR area. All p-values are from Wald $\chi^2$ tests jointly over all included lag terms. Bold indicates $p<0.05$. Key findings from this sweep support the main-text claims: (i) ALPM Granger-predicts MR in VFMR primarily at lags 1–4 and at lag 7, with isolated additional hits at lags 13 and 19; (ii) ALPM is non-predictive at every lag in AFMR; (iii) MR predicts ALPM at every lag in AFMR (strongly asymmetric reverse direction), with lag 1 $p=0.016$ and lags 2–20 all $p<0.001$; (iv) MR does not predict ALPM at any lag in VFMR (minimum $p=0.061$ at lag 9). AFMR = atrial functional mitral regurgitation; VFMR = ventricular functional mitral regurgitation; ALPM = anterolateral papillary muscle; LV = left ventricle; LA = left atrium; MR = mitral regurgitation.*

**Supplementary Table S2.** Intraobserver Reproducibility — Bland–Altman Analysis

| **Variable** | **N** | **ICC** | **95% CI** | **Mean bias** | **SD diff** | **LoA lower** | **LoA upper** | **CV%** |
|---|---|---|---|---|---|---|---|---|
| MR area ($cm^2$) | 70 | 0.986 | 0.978–0.991 | +0.003 | 0.097 | −0.187 | 0.193 | 6.3 |
| LA volume (ml) | 70 | 0.981 | 0.970–0.988 | +0.151 | 2.875 | −5.484 | 5.787 | 3.2 |
| LV volume (ml) | 70 | 0.971 | 0.959–0.984 | −2.349 | 5.330 | −12.796 | 8.098 | 8.6 |
| ALPM length (mm) | 70 | 0.890 | 0.827–0.929 | +0.009 | 1.254 | −2.449 | 2.467 | 5.0 |

*Intraobserver reproducibility assessed in 7 patients (70 paired systolic-frame observations per variable). ICC = intraclass correlation coefficient, two-way mixed effects model, absolute agreement, single measures (ICC 2,1). Mean bias = mean of (repeat − original). SD diff = standard deviation of paired differences. LoA = limits of agreement (mean bias ± 1.96 × SD; Bland–Altman method). CV% = coefficient of variation, calculated as (SD of differences / grand mean) × 100. ALPM = anterolateral papillary muscle; LA = left atrium; LV = left ventricle; MR = mitral regurgitation.*
